# Maintenance Electroconvulsive Therapy and hospital re-admission days among patients with mental illness in Malaysia

**DOI:** 10.1101/2020.09.20.20197079

**Authors:** Benedict Francis, Ng Yit Han, Julian Wong Joon Ip, Ling Shiau Thin, Jesjeet Singh Gill, Koh Ong Hui, Chiara Francine Petrus, Ahmad Hatim Sulaiman

## Abstract

**Background:** Relapse prevention strategies are important as part of optimal patient care. As such, maintenance electroconvulsive therapy (mECT) is an important treatment modality to keep patients in remission longer. However, the practice of mECT in Malaysia, specifically and Asia generally, is still scarce.

**Aims:** Our study aimed to explore the characteristics of patients receiving maintenance ECT (mECT) and further investigate whether this treatment modality reduces re-admission days in patients with severe mental illness.

**Methods:** A retrospective chart review study design was employed. The medical records of 22 patients followed up at University Malaya Medical Centre, Malaysia were analysed with regards to the outcome measures, which was days of re-admission post mECT, Potential confounders were controlled for via stratification analysis.

**Results:** There was a significant reduction in re-admission days post mECT (p<0.001, r:0.85) across all the variables analysed. The variable with the biggest effect size were patients younger than 60 years old (p:0.01, r:0.70), followed by medication with polypharmacy (p:0.002, r: 0.65). The magnitude of reduction in re-admissions was greater in the schizophrenia spectrum group compared to the affective disorders group (r: 0.64 vs. 0.57).

**Conclusion:** Our study provides data regarding the efficacy of mECT in significantly reducing days of re-admission in patients with treatment resistant schizophrenia spectrum illness and affective disorders. As widespread usage of mECT is still lacking in Asia, our results is encouraging for more practitioners to prescribe mECT for their patients in order to reduce rates of hospital re-admission.

## INTRODUCTION

At the dawn of the new decade of the 21^st^ century, Electroconvulsive Therapy (ECT) remains the safest and most used somatic therapy in the field of neuropsychiatry since its inception in 1938(1). In fact, ECT has been showed to be effective in a myriad of psychiatric diagnoses, particularly schizophrenia and affective disorders (2, 3). As relapse prevention and prolonging remission is the goal of holistic psychiatric care, it is imperative that effective strategies be employed early. Research has shown that in treatment resistant illness, adjunct maintenance ECT (mECT) with psychotropic medication led to superior rates of remission (4). As such, it is the treatment modality of choice in severe and treatment resistant mental illnesses. Continuous ECT (cECT) refers to treatments given at regular intervals of one week to one month, within the first 6 months after an acute course of ECT has been done. Maintenance ECT (mECT), on the other hand, refers to the same, regularly spaced treatment given after the initial 6-month period (5).

Upon recovery with acute courses of ECT, it is imperative to consider cECT and mECT to reduce rates of relapses. Indeed, several authors have demonstrated the superiority of continuous ECT compared to placebo and other active treatments in reducing long term relapse rates (6-8). In particular, long term mECT is associated with better outcomes with regards to efficacy, side effect profile and long-term tolerability (9).

Although there has been emerging evidence from South East Asian countries with regards to the efficacy of mECT in relapse prevention, its usage is still lacking. This pattern is surprising, given that ECT itself is not an uncommon practice among Asian psychiatrists (10). To the authors’ knowledge, there has been no such prior study done in Malaysia. The present study aimed to investigate the relationship between mECT and hospital re-admission days in a Malaysian setting.

## METHODOLOGY

### Study design and setting

This study was conducted via a retrospective chart review design. The medical records of 156 patients that underwent mECT in University Malaya Medical Centre (UMMC) from January 2010 – December 2019 were obtained and reviewed. UMMC is a tertiary hospital and a referral centre for complex psychiatric cases. The detailed flowchart of patient selection is shown in Figure 1.

**Figure 1:**
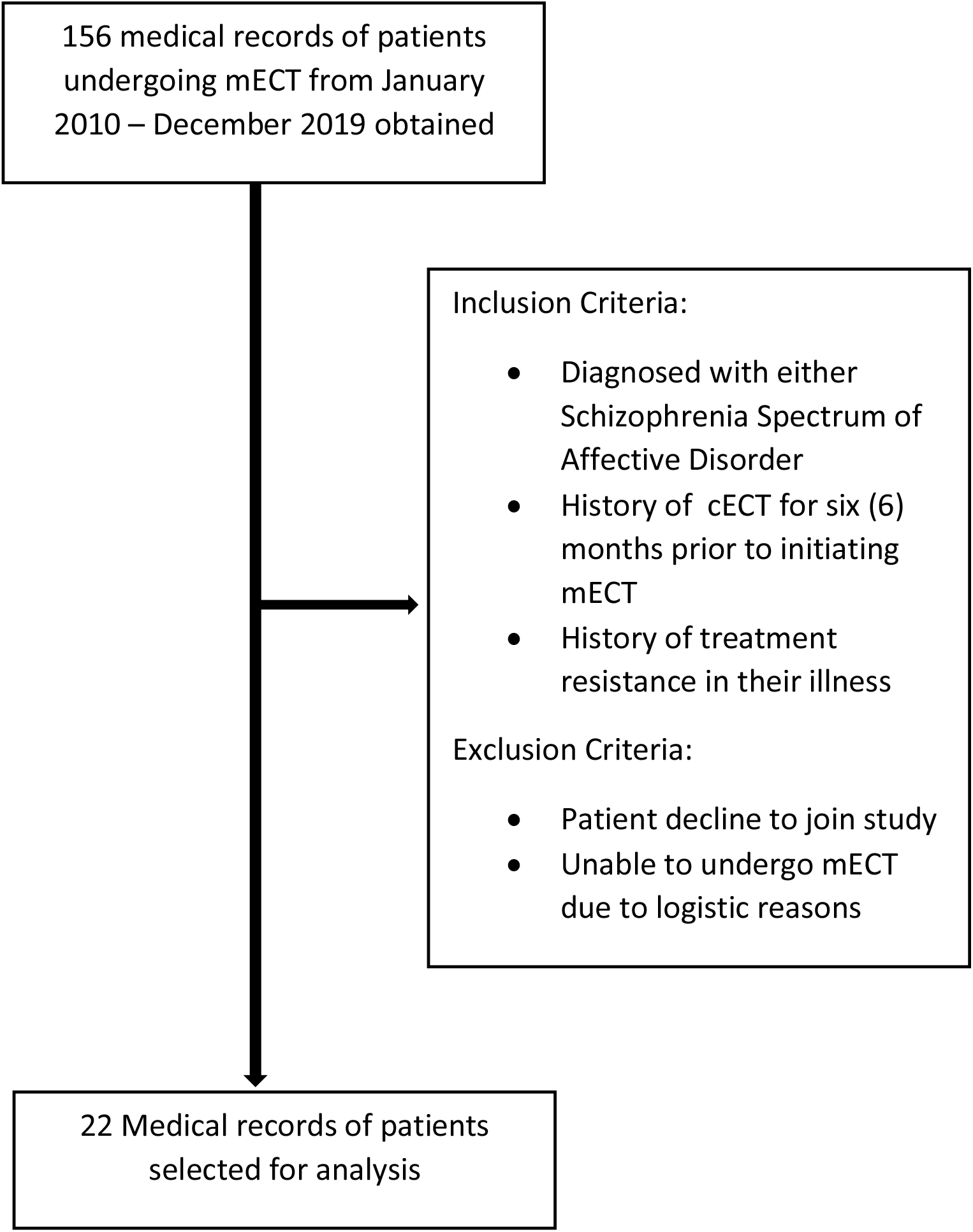
Flowchart of patient selection. * mECT: Maintenance electroconvulsive therapy cECT: Continuous electroconvulsive therapy

For this audit, patients’ socio-demographic and clinical data were collected. Re-admission was defined as any hospitalization due to a relapse in their psychiatric illness post mECT. ECT was administered using the Thymatron^R^ System IV, with bitemporal lead placement. All of the patients suffered from either schizophrenia-spectrum illness (schizophrenia and schizoaffective disorders), or affective disorders (major depressive disorder or bipolar disorders). The following inclusion and exclusion criteria were applied: Inclusion criteria:

a. Had undergone a course of cECT for 6 months prior to initiating mECT.
b. Diagnosed with either a schizophrenia spectrum or affective disorder
c. History of treatment resistance in their illness. Treatment resistance was defined as non-response to at least 2 appropriate medications.

Exclusion criteria were as follows:

a. Patients declined to join study.
b. Unable to receive mECT due to logistic reasons.

For this study, the following definition of mECT was adopted: the administration of additional ECTs after the patient had obtained clinically significant remission. Treatment resistance was defined as a non-response to 2 different pharmacologic treatments from different classes, after an adequate duration and dosage. Remission was defined as Clinical Global Impression-Severity (CGI-S) rating of either 1 or 2 after 2 consecutive assessments(11).

After applying the criteria above, the medical records were narrowed down to 22 patients, who fulfilled the inclusion and did not fulfil the exclusion criteria.

### Timeframe

The 2010-2019 timeframe was selected to allow adequate comparison between pre and post mECT re-admission days. Days of re-admission pre and post mECT were taken within an equal time frame. For example, if the patient underwent 1 year of mECT, the period of re-admisions 1 year before and 1 year after the mECT were taken into account.

### Statistical Analysis

The median with inter-quartile range was used to represent continuous variables, as the data was not normally distributed. Categorical typed variables were represented by frequency and percentage. A normality test was conducted to examine the mean difference in days of hospitalization (pre and post maintenance ECT). In view of the skewed distribution of the data, the Wilcoxon-Sign Ranked test was used instead of the paired t-test. The data was presented as interquartile range and median, together with the p value and effect size, r.

An overall median difference in terms of days of hospitalization baseline versus post mECT was calculated. As the data was not normally distributed, the Wilcoxon-Sign ranked test is used instead of paired t-test. Significant difference between days of hospitalization before and after treatment is indicated by p < 0.05.

Potential demographic and clinical confounders to the outcome measure were controlled for via stratification analysis. The stratification analysis was applied to ensure that the treatment did indeed affect the days of hospitalization after considering these third variables. Statistical Package for Social Science version 25.0 (IBM Corp.,Armonk,NY,USA) was used to analyse the data.

## RESULTS

Table 1 shows the socio-demographic information. A total of 22 patients were analysed in this study, with the majority of them females (59.1%). Most patients were of the younger age group, with 63.6% of them younger than 60 years old. A majority of them suffered from schizophrenia spectrum disorders compared to affective disorders (54.5 % vs 45.5 %). The median duration of illness of the cohort studied were 8 years, where else in term of number of ECTs done, half of them were within 40 mECTs. A slight majority of patients had comorbid medical illness (54.5 %).

**Table 1:**
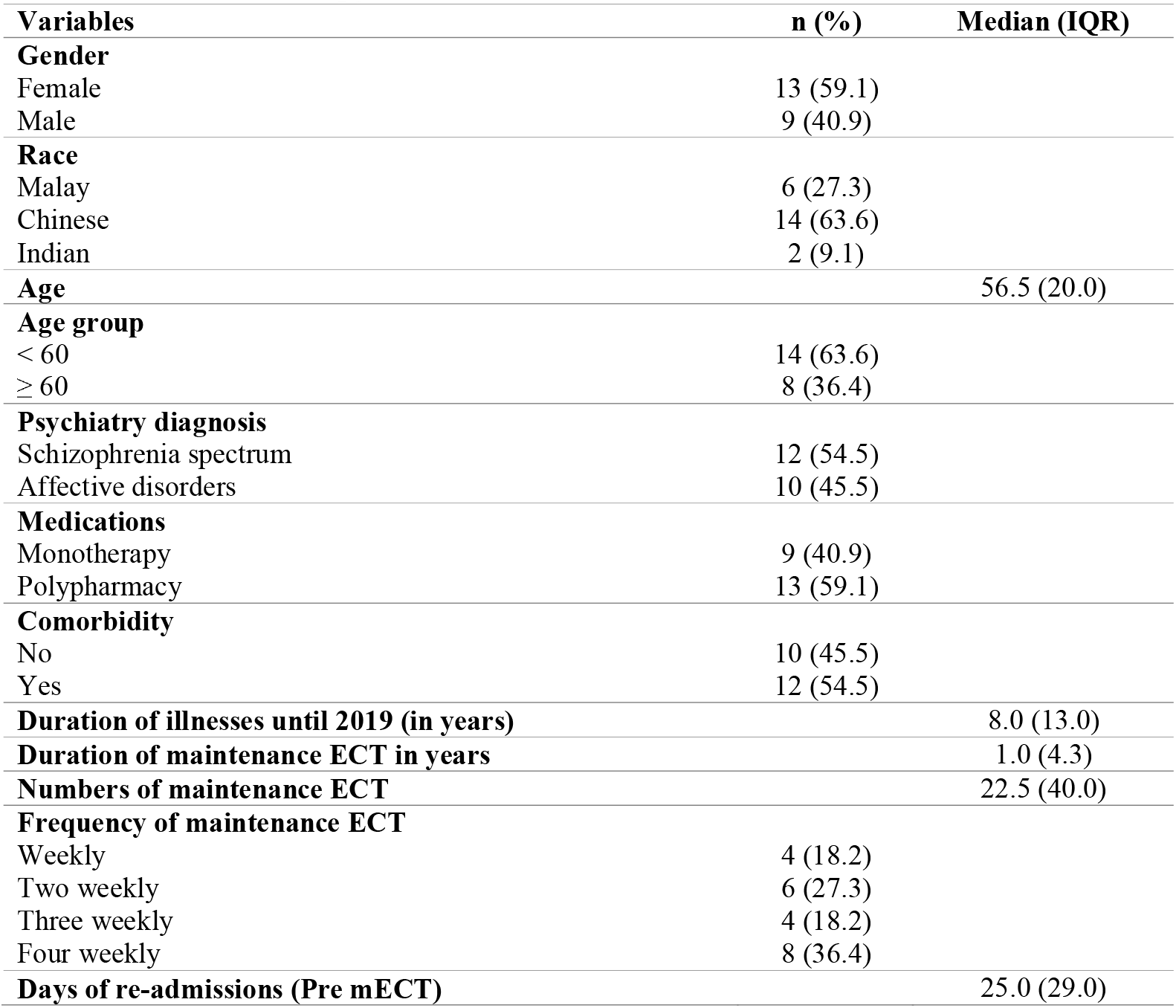
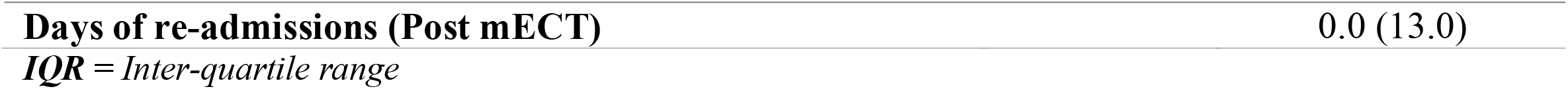
Demographic profiles and clinical characteristics of the patients underwent maintenance ECT therapy (n=22)

**Table 2:**
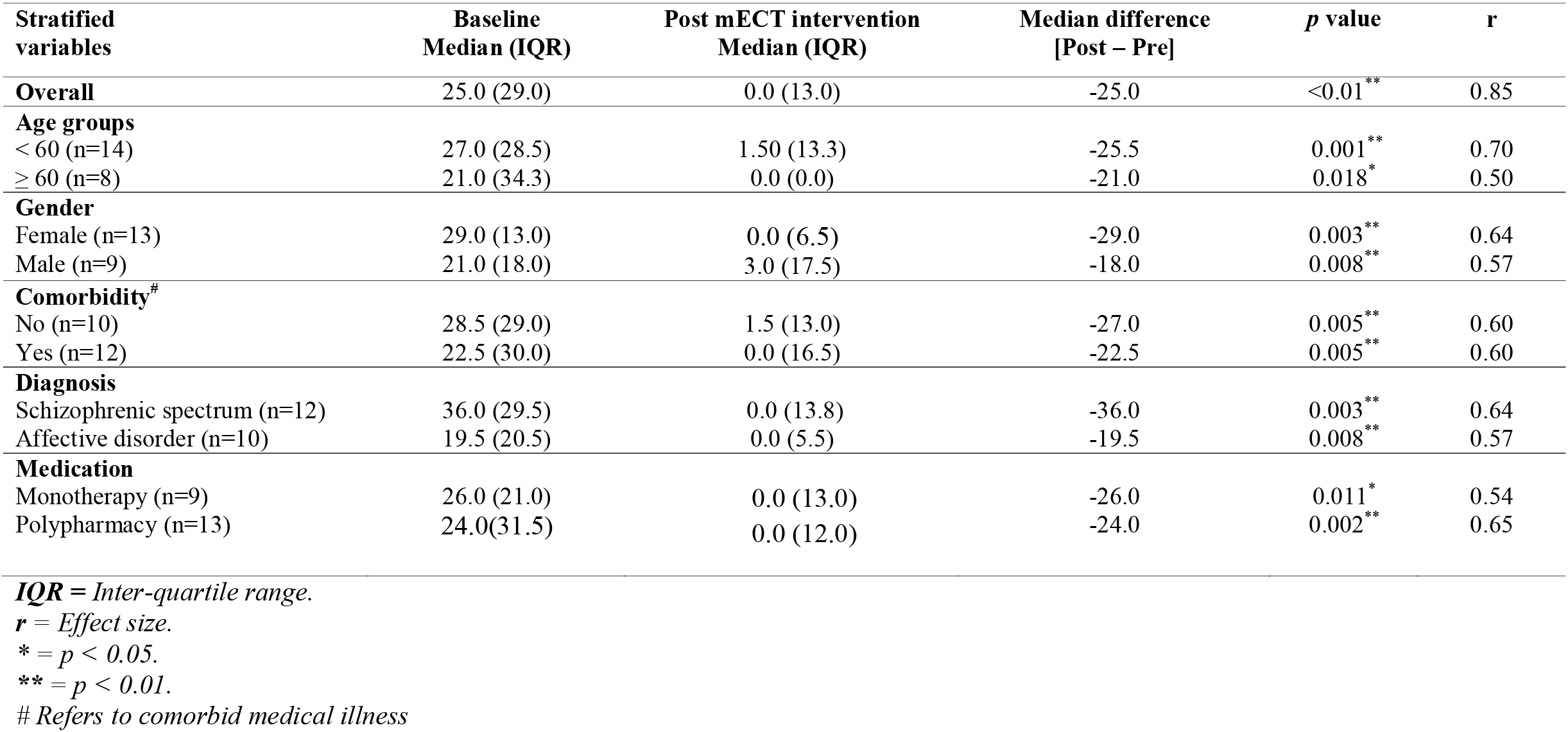
Comparison of median differences between number of days of re-admission baseline and post maintenance ECT (mECT) treatment among patients (n=22)

| Stratified variables | Baseline Median (IQR) | Post mECT intervention Median (IQR) | Median difference [Post – Pre] | *p* value | *r* |
| --- | --- | --- | --- | --- | --- |
| **Overall** | 25.0 (29.0) | 0.0 (13.0) | -25.0 | <0.01** | 0.85 |
| **Age groups** |  |  |  |  |  |
| < 60 (n=14) | 27.0 (28.5) | 1.50 (13.3) | -25.5 | 0.001** | 0.70 |
| ≥ 60 (n=8) | 21.0 (34.3) | 0.0 (0.0) | -21.0 | 0.018* | 0.50 |
| **Gender** |  |  |  |  |  |
| Female (n=13) | 29.0 (13.0) | 0.0 (6.5) | -29.0 | 0.003** | 0.64 |
| Male (n=9) | 21.0 (18.0) | 3.0 (17.5) | -18.0 | 0.008** | 0.57 |
| **Comorbidity<sup>#</sup>** |  |  |  |  |  |
| No (n=10) | 28.5 (29.0) | 1.5 (13.0) | -27.0 | 0.005** | 0.60 |
| Yes (n=12) | 22.5 (30.0) | 0.0 (16.5) | -22.5 | 0.005** | 0.60 |
| **Diagnosis** |  |  |  |  |  |
| Schizophrenic spectrum (n=12) | 36.0 (29.5) | 0.0 (13.8) | -36.0 | 0.003** | 0.64 |
| Affective disorder (n=10) | 19.5 (20.5) | 0.0 (5.5) | -19.5 | 0.008** | 0.57 |
| **Medication** |  |  |  |  |  |
| Monotherapy (n=9) | 26.0 (21.0) | 0.0 (13.0) | -26.0 | 0.011* | 0.54 |
| Polypharmacy (n=13) | 24.0(31.5) | 0.0 (12.0) | -24.0 | 0.002** | 0.65 |

***IQR*** = Inter-quartile range.
***r*** = Effect size.
\* = $p < 0.05$ .
\*\* = $p < 0.01$ .

The median difference in Interquartile Range (IQR) between days of re-admission pre and post mECT was highly significant (r:0.85, P<0.01, median difference: −0.25). Our results showed that 50 % of patients post mECT were within 13 days of readmission (IQR:13) compared to within 29 days in the pre mECT group (IQR:29). The variable with the biggest effect size were patients younger than 60 years old (p:0.001, r:0.70), followed by medication with polypharmacy (p:0.002, r: 0.65). The magnitude of reduction in re-admissions was greater in the schizophrenia spectrum group compared to the affective disorders group (r: 0.64 vs. 0.57).

Out of the 22 patients that were included in this chart review, none dropped out due to adverse effects of mECT. 7 patients complained of post procedure headaches, and 3 patients complained of transient nausea. There were no complaints of prolonged cognition impairment or confusion.

## DISCUSSION

### Main findings

Relapse prevention and prolonged remission leading up to functional recovery is the mainstay of psychiatric interventions. In patients who have achieved remission in their illness after an acute course of ECT, the need for continuous courses of ECT as a form of relapse prevention arises (12). Although there have been previous Asian studies on ECT and relapse prevention (13-15), there is a paucity of studies looking into usage of mECT in the South East Asian region (16). Our study provides important national data to reinforce the notion that mECT reduces days of re-admission.

Our data showed a very significant reduction in re-admission after patients underwent mECT. Among these patients, those who were aged less than 60 years old showed the most improvement, similar with the results obtained by O’Connor and colleagues (17). Younger patients are often more robust, have lesser comorbid illness and thus respond better to mECT. Older patients, in contrast, are a unique population as they often have other confounding factors to their recovery such as functional disability, social isolation and pain (18). The fact that these patients may be lonelier than their younger counterparts plays a role as loneliness is associated with increased healthcare utilization, and thus increased disease burden (19). These factors synergize with other unmet healthcare needs, resulting in increased morbidity (20).

The most common indication for mECT in our cohort was schizophrenia spectrum illnesses. This result may reflect an Asian preference, as a similar result was also reported from a survey conducted among psychiatric facilities across Asia (21). In fact, this line of thought is congruent with recent evidence proving mECT combined with maintenance antipsychotics are superior that either treatment alone in managing treatment resistant schizophrenia (22-24). In contrast to the common notion that mECT works better in affective disorders (25, 26), our data showed higher gradient of improvement for patients with treatment resistant schizophrenia spectrum illness compared to affective disorders. This result is a unique finding in our population and warrants further exploration. One possible reason for this finding could be that patients in our study population with affective disorders had stronger and more persistent environmental stressors that perpetuated their illness even after discharge post mECT. This led them to have more severe illness and thus improve with a lesser quantum.

### Limitations

It is important to state the limitations of this study. The relatively small sample size may hinder generalizability of the results. However, most studies done on mECT suffer from relatively small sample sizes, partly due to the specialised and unique nature of the treatment modality (23). The retrospective chart review design made determination of cause and effect not possible, and the collation of data relied solely on the accuracy of recorded data. Although efforts were made to control for confounders by means of stratification analysis of the variables, a prospective design would have controlled for bias and confounders better.

### Implications

There seems to be a sense of reticence to wider usage of mECT globally among patients who do not respond to pharmacological treatment (27). Part of the problem seems to be stigma against widespread ECT usage, coupled with an acute lack of psychoeducation with regards to its efficacy and safety even amongst prescribers. Our study showed that mECT significantly reduced re-admission days in patients with severe mental illness. In addition, patients with schizophrenia spectrum illnesses had a greater magnitude of change in re-admission days compared to those with affective disorders. Thus, mECT should be considered in the maintenance regime of these patients, as it is safe and highly efficacious. Future research should focus on prospective, randomized and blinded designs in order to further elucidate the relationship between mECT and readmissions days in a controlled fashion, particularly in Asia.

## Data Availability

Nil

## Acknowledgement

The authors would like to acknowledge Dr Stephen Thevananthan Jambunathan, whose vision and zeal inspired this work.

